# Analysis of COVID-19 spread in South Korea using the SIR model with time-dependent parameters and deep learning

**DOI:** 10.1101/2020.04.13.20063412

**Authors:** Hyeontae Jo, Hwijae Son, Hyung Ju Hwang, Se Young Jung

## Abstract

Mathematical modeling is a process aimed at finding a mathematical description of a system and translating it into a relational expression. When a system is continuously changing over time (e.g., infectious diseases) differential equations, which may include parameters, are used for modeling the system. The process of finding those parameters that best fit the given data from the system is called an inverse problem. This study aims at analyzing the novel coronavirus infection (COVID-19) spread in South Korea using the susceptible-infected-recovered (SIR) model. We collect the data from Korea Centers for Disease Control & Prevention (KCDC). We assume that each parameter in the SIR model is a function of time so that we can compute important parameters, such as the basic reproduction number (R0), more delicately. Using neural networks, we propose a method to find the best time-varying parameters and the solution for the model simultaneously. Moreover, using time-dependent parameters, we find that traditional numerical algorithms, such as the Runge-Kutta methods, can successfully approximate the SIR model while fitting the COVID-19 data, thus modeling the propagation patterns of COVID-19 more precisely.

## 1. Introduction

Similar to the 1918 influenza pandemic (known as the “Spanish Flu”) more than 100 years ago, the coronavirus disease 2019 (COVID-19) pandemic has been a major shock to the whole world. At the end of World War I, the 1918 influenza pandemic wreaked havoc on the whole world, killing more than 40 million people, more than 2% of the then world’s population of 1.8 billion people. It is estimated that 16 million people died in India, which suffered the most damage, while in the US, the pandemic killed 5.2% of the entire population [1]. Additionally, it had a catastrophic impact on the world economy. For example, manufacturing production fell by an average of 18% in the US, since many people died or became sick, resulting in a devastating loss in the workforce, while the fears of infection caused a decrease in consumption and production. During the outbreak, preventive measures such as social distancing and wearing masks were recommended to curb the spread of the virus. However, the 1918 influenza pandemic exhibited an unusual bimodal peak in the US, lasting for almost two years. Thus, by analogy, we can infer that decreases in infection rates of COVID-19 do not guarantee continuity of the trend.

Nevertheless, the current status of the COVID-19 pandemic is worse than that of the 1918 influenza pandemic due to the increasingly entangled transportation network connecting the world and the global interdependency between economies that developed over decades of globalization. Even countries that have successfully imposed quarantine measures early, such as Taiwan and Hong Kong, cannot rule out the possibility of a COVID-19 re-outbreak. Therefore, governments and academia need to collect and analyze all COVID-19-related data using big data technologies in real time to monitor the pandemic by region and time quantitatively. Moreover, the effects of environmental changes and policies should be evaluated promptly so that appropriate measures can be provided immediately according to the results.

The susceptible-infected-recovered (SIR) model is one of the simplest and powerful models that can be used to mathematically model infectious diseases. Together with its three main variables (S, I, R) the model is governed by two parameters that represent the transition rates between two variables, *β* and *γ*.

Recently, there have been numerous works analyzing COVID-19. Some studies regarded the parameters as constants. For instance, [2] proposed a conceptual model of COVID-19 that includes individual behavioral reaction and government actions, while [3] reviewed the basic reproduction number (R0), one of the most important indicators, of COVID-19. Other studies, such as [4], divided the phase manually and regarded the parameters as time-varying piece-wise constants. Moreover, [5, 6] considered the parameters as functions of time and proposed methods to approximate the time-varying parameters. Additionally, [7, 8] have shown that neural networks (NNs) with proper loss functions represent a powerful tool for solving forward-inverse problems. Most previous studies considered the parameters constants because of the complexity of the model. However, it has been reported that constant parameters cannot model the actual data precisely.

This study analyzes the spread of COVID-19 in South Korea, using the SIR model. We regard the modeling problem as a forward-inverse problem, and approximate each variable (S, I, R) and parameter (*β, γ*) in the model using deep learning. Moreover, to overcome the shortcomings of previous studies, we consider the parameters as functions of time, which allows us to compute the infection rate, recovery rate, and the time-dependent reproduction number, *R*_*T D*_. This approach is more interpretable, since *β*(*t*), *γ*(*t*), *R*_*TD*_ can be obtained as functions of time, as well as the overall dynamics of the actual data. Additionally, unlike other models, such as the growth model, we do not assume any distribution type for the modeling. In the traditional growth model, one considers the growth rate as a piecewise constant function to compute the effective reproduction number. However, this assumption is not realistic in many cases, as the reproduction number could dramatically change. In contrast, the time-dependent nature of our model makes it an appropriate solution for such problems. Furthermore, we provide numerical simulation results that guarantee the convergence of our NN approach. Finally, our methodology is applicable to many areas dealing with differential equations, and easy to implement without a deep understanding of the model.

The time-dependent reproduction number, *R*_*TD*_ is a metric of a pathogen’s transmissibility [9]. *R*_*T D*_ is an important measure for assessing whether current efforts are effective or additional intervention is needed [10]. This study establishes a more accurate reproduction number prediction model by adopting a deep learning approach for the SIR model with time-dependent parameters to evaluate the effectiveness of intervention in curbing the spread of COVID-19 in South Korea.

The remainder of this paper is organized as follows. Section 2 provides information about the data and the data sources. Section 3 gives a precise definition of the SIR model. Section 4 explains the deep NN (DNN) approach. Section 5 presents the simulation results using DNNs. Finally, section 6 discusses the findings and summarizes the conclusions.

## 2. Data

We collect the data from Korea Centers for Disease Control & Prevention (KCDC). The data consist of the cumulative number of tested people (*T*), confirmed cases (*I*, or *I*_*pos*_), negative cases (*I*_*neg*_), and recovered or deceased cases (*R*) from February 7, 2020 to March 30, 2020 for South Korea and from March 5, 2020 to March 30, 2020 for the administrative provinces of Seoul, Busan, and Daegu.

## 3. SIR Model

We first introduce the SIR model. For a fixed time *t* ≥ 0, let *S*(*t*), *I*(*t*), *R*(*t*), and *N* (*t*) denote the numbers of susceptible, infected, and recovered (or removed) populations, and the sum of these three variables, respectively. Thus, we can write the SIR model as

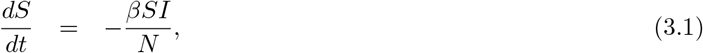

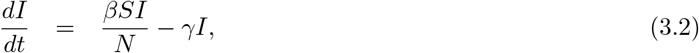

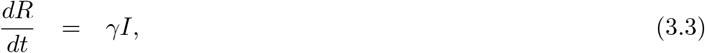

where *β >* 0 and 0 *< γ <* 1 denote the infection and the recovery rates, respectively. In general, we set the initial condition *S*(0), *I*(0), and *R*(0) as the first observation data, and for the analysis, we assume that the total number of population *N* is time-invariant, that is,

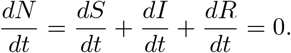

In this study, We applied a scaled SIR model (divided by *N* for each variable *S, I*, and *R*) and time-varying parameters (*β, γ*), with the final SIR model being as follows:

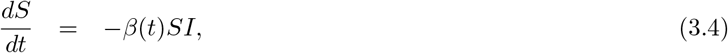

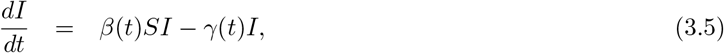

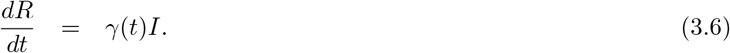

For the observation data, we set 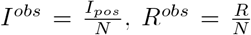, and *S*^*obs*^ = 1 *− I*^*obs*^ *− R*^*obs*^. The time-dependent reproduction number, *R*_*T D*_(*t*), is defined by *R*_*T D*_(*t*) = *β*(*t*)*/γ*(*t*).

## 4. Deep Learning: DNNs

Deep learning is a non-linear function approximation method that uses DNNs. A DNN consists of one input layer, one output layer, and several hidden layers, with the adjacent layers connected by an affine transformation followed by a non-linear activation function. NNs were first introduced in [11]. Then, Cybenko [12] proved that an NN with a single hidden layer can approximate any continuous function under some conditions on the activation function. Additionally, Hornik-Stinchcombe-White [13] showed that a multilayer feedforward NN with a sigmoid activation function can approximate any measurable function. Later, Li [14] proved that an NN with one hidden layer can simultaneously approximate any measurable function and its partial derivatives on a compact set.

Specifically, let 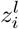 be the *i*^*th*^ neuron in the *l*^*th*^ layer. Then,

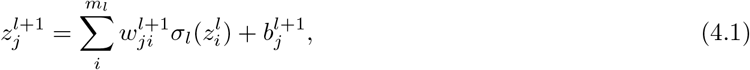

where

- *m*_*l*_: the number of neurons in *l*^*th*^ layer
- *σ*_*l*_ the activation function in *l*^*th*^ layer
- 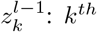 neuron in (*l −* 1)^*th*^ layer
- 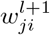: the weights between *i*^*th*^ neuron in *l*^*th*^ layer and *j*^*th*^ neuron in (*l* + 1)^*th*^ layer
- 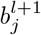: the bias of *j*^*th*^ neuron in *l*^*th*^ layer

We constructed five NN models for *S, I, R, β*, and *γ*, denoted by *S*_*net*_, *I*_*net*_, *R*_*net*_, *β*_*net*_, and *γ*_*net*_, respectively. The concrete model structures are presented in Figure 6. We applied similar training methods as introduced in [8] and [7].

**Figure 1:**
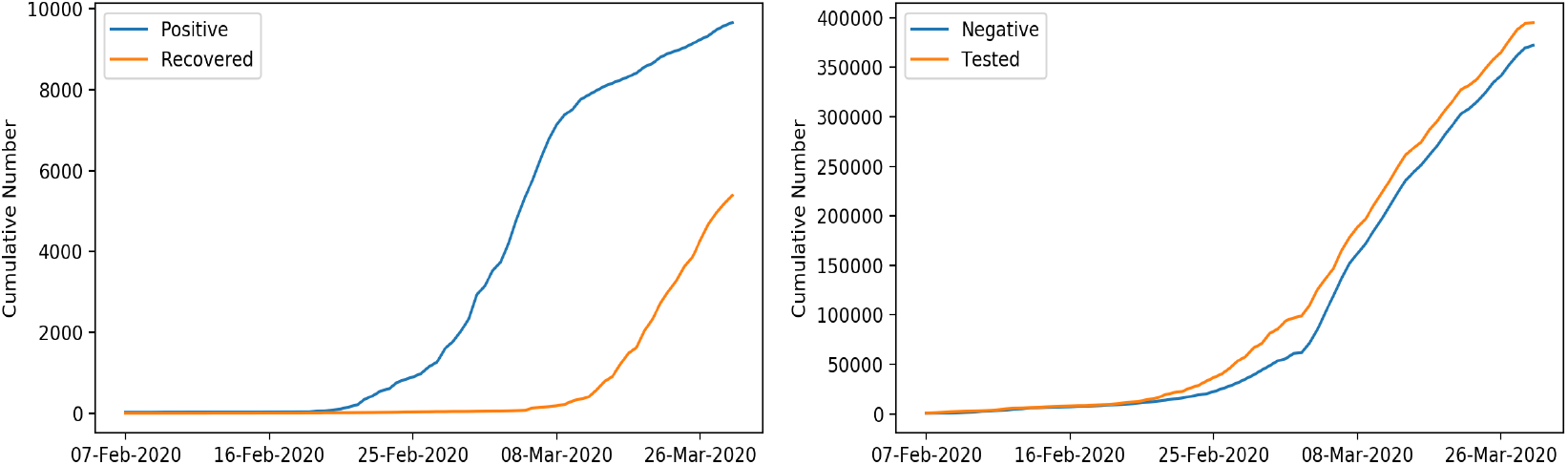
Left: Cumulative number of infected cases and recovered cases, Right: Cumulative number of negative cases and tested people

**Figure 2:**
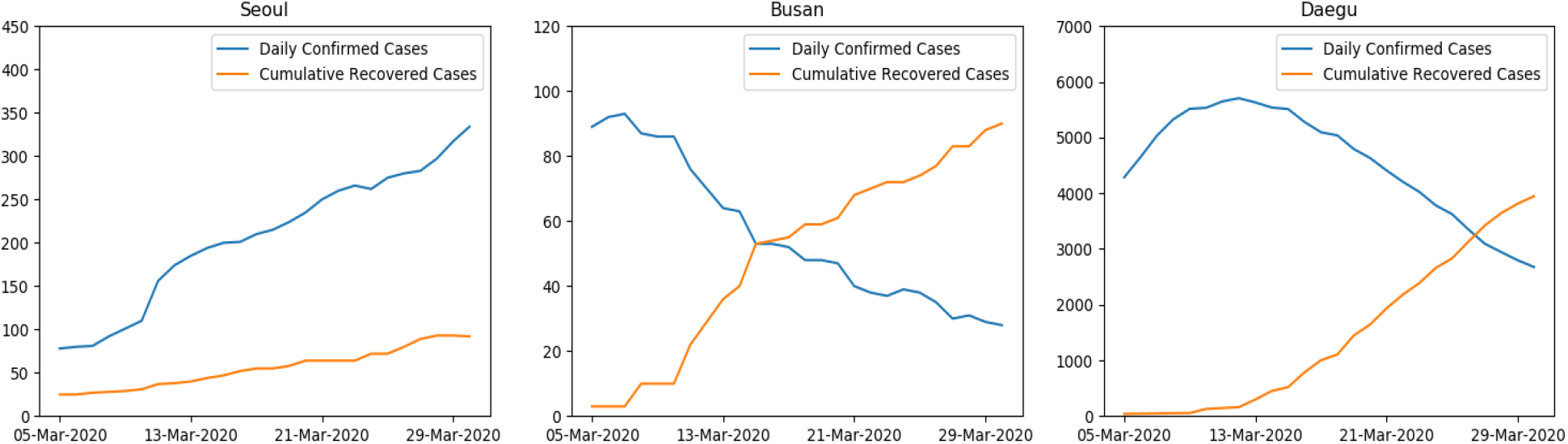
Daily number of confirmed cases and cumulative number of recovered cases for Seoul, Busan, and Daegu

**Figure 3:**
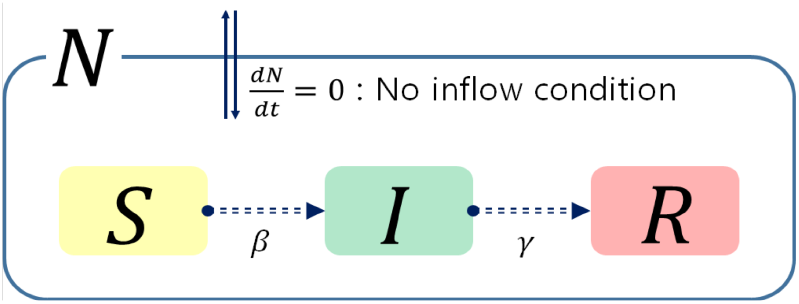
SIR model

**Figure 4:** Graphical description of the SIR models

**Figure 5:**
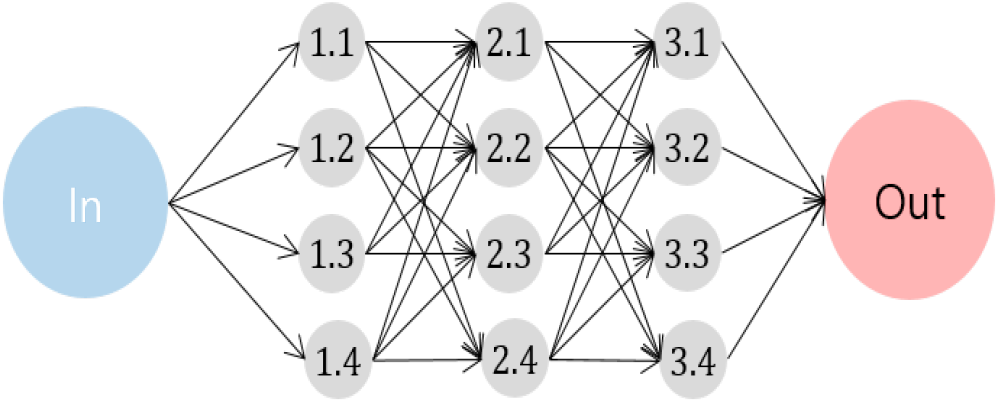
Pictorial description of the DNN architecture. The above network has three hidden layers, and each hidden layer consists of four neurons (nodes). Pairs (*x, y*) in nodes mean *y*^*th*^ neuron in *x*^*th*^ layer.

**Figure 6:**
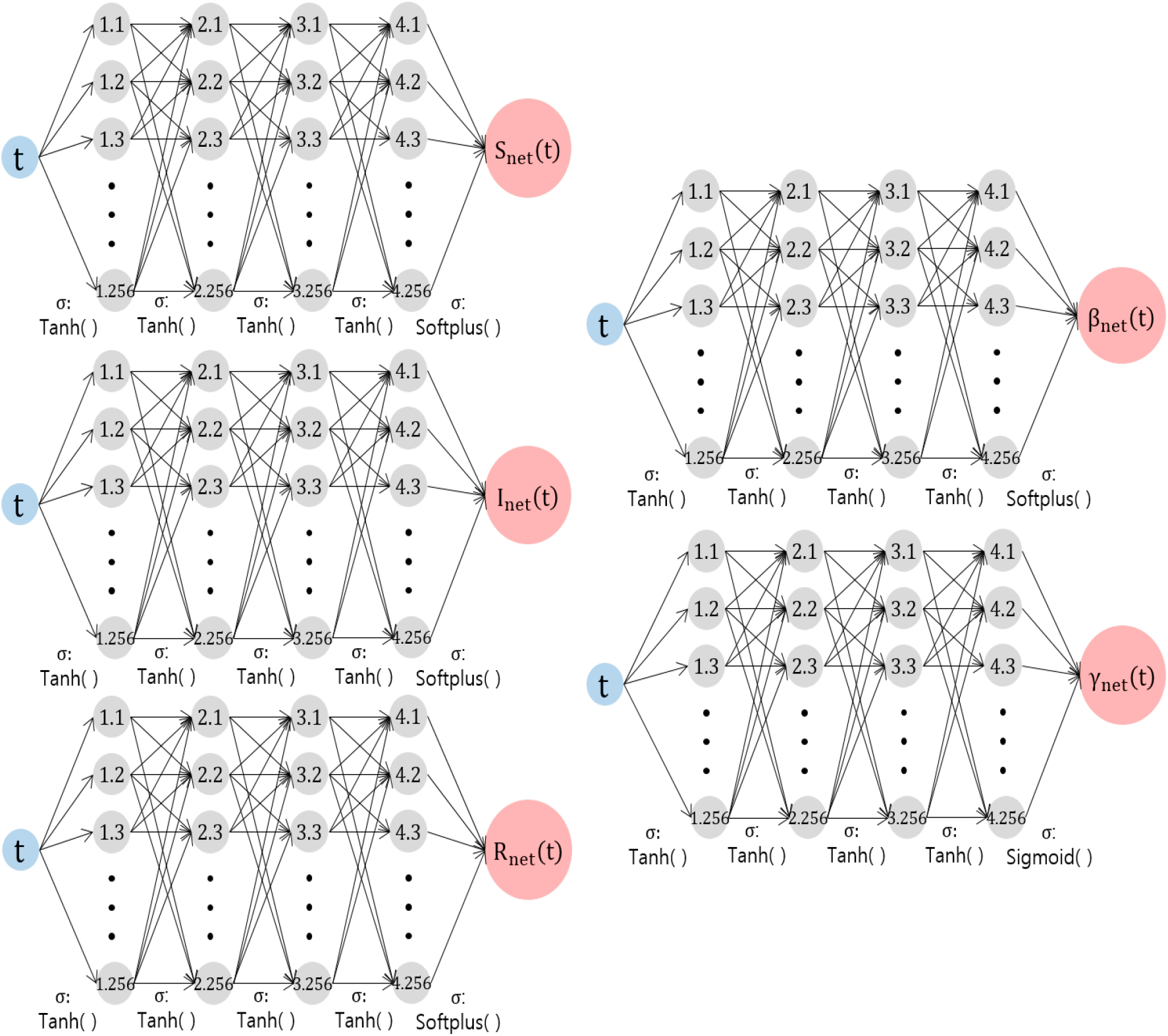
Forward-Inverse SIR model networks. Each network has one input node (time *t*), one output node (value), four hidden layers, and 256 nodes in each hidden layer. The hyperbolic tangent *tanh*(*x*) is used in the activation functions except for the last layers. The Softplus and Sigmoid functions are used in the last hidden layer to meet the constraints *S, I, R, β* > 0, and 0 < *γ* < 1.

## 5. Simulation Result

We conducted simulations for three provinces (Seoul, Busan, and Daegu) and South Korea, using the total population numbers *N* as shown in Table 1.

**Table 1:**
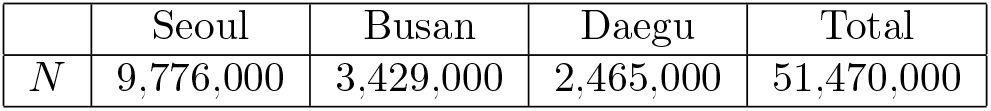
Total population numbers for each province and South Korea.

In this section, we provide the simulation results for five NN models: *S*_*net*_, *I*_*net*_, *R*_*net*_, *β*_*net*_, and *γ*_*net*_ and the corresponding relative errors 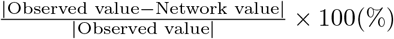 for *S, I, R*. For a more accurate evaluation of the model parameters, we also provide numerical solutions (Runge-Kutta of order 4, (RK4)) using the estimated parameters. For the RK4 method, we set *h* = 10^*−*3^ (step size), with 26 observations used for Seoul, Busan, and Daegu, and 77 observations for South Korea. We remark that the observations are available on the KCDC homepage, http://www.cdc.go.kr/. In Figure 7 - 10, Red lines denote *S*_*net*_, *I*_*net*_, *R*_*net*_ values for each graph, Green lines denote the observations, and Blue lines denote the RK4 results with the parameter NN *β*_*net*_, *γ*_*net*_. In Figure 11 - 14, we provide the estimated time-dependent parameters *β*, and *γ* on the left, and the time-dependent reproduction number *R*_*T D*_ on the right. Next, we summarize the overall trend by analyzing *R*_*T D*_(*t*). First, at *t* = 0 in South Korea, *R*_*T D*_ = 1.0610 implies the spread of COVID-19, see Figure 11. Starting from 18, February, *R*_*T D*_(*t*) increased dramatically (*t* = 11), and reached its peak (*R*_*T D*_(*t*) = 124.7610) on 27, February (*t* = 20.3). After 13 March (*t* = 34.6), *R*_*T D*_ fell below 1 again, signaling a decreasing trend in the spread of COVID-19 from an epidemiological viewpoint. However, it started increasing again from 18, March (*t* = 39.6), pointing out that the containment of COVID-19 cannot be realized without achieving herd immunity or developing therapeutics. From 27, February to 30, March, the average values of *β* and *γ* were 0.1656, 0.0253, respectively. In the second case, Seoul, until 9, March (*t* = 4), *β* reached 0.2306, while *gamma* reached only 0.0192, resulting in a maximum value of 12.0405 for *R*_*T D*_ in this period. After 16 March (*t* = 11), *R*_*T D*_ decreased to 3.1244 but increased again reaching 3.8255 on 30 March (*t* = 25, see Figure 12, right). This indicates that no effective control of the spread of COVID-19 has been achieved. The average values of *β* and *γ* were 0.0705, 0.0140, respectively. In the third case, Busan, on 5, March (*t* = 0), at the beginning of the observation, *β* was 0.1300 (Figure 9, left bottom,) while *R*_*T D*_ was 156.7965. This is because *R*(*t*), the recovery group, did not change in the initial stage, whereas *γ* was estimated to be 0.0008 due to the constraint *γ >* 0. On 8, March (*t* = 3), *R*_*T D*_ was 0.0908 because of the change in *R*(*t*), reaching 0.5401 on 30, March (*t* = 25). We also provide two *R*_*T D*_ plots by dividing the observation period into two periods in Figure 13. The average values of *β* and *γ* were 0.0253, 0.0670, respectively. In the final case, Daegu, similar to Busan, *R*_*T D*_ was 521.9075 at the beginning of the observation on 5, March (*t* = 0). After 11, March (*t* = 6), the recovery rate *γ* began to increase faster than the infection rate *β*, with the *R*_*T D*_ having the lowest value 0.1224 on 24, March (*t* = 19). After 24, March, *R*_*T D*_ increased reaching 0.2409 on 30, March (*t* = 25), see Figure 14. The average values of *β* and *γ* were 0.0191, 0.0387, respectively.

**Figure 7:**
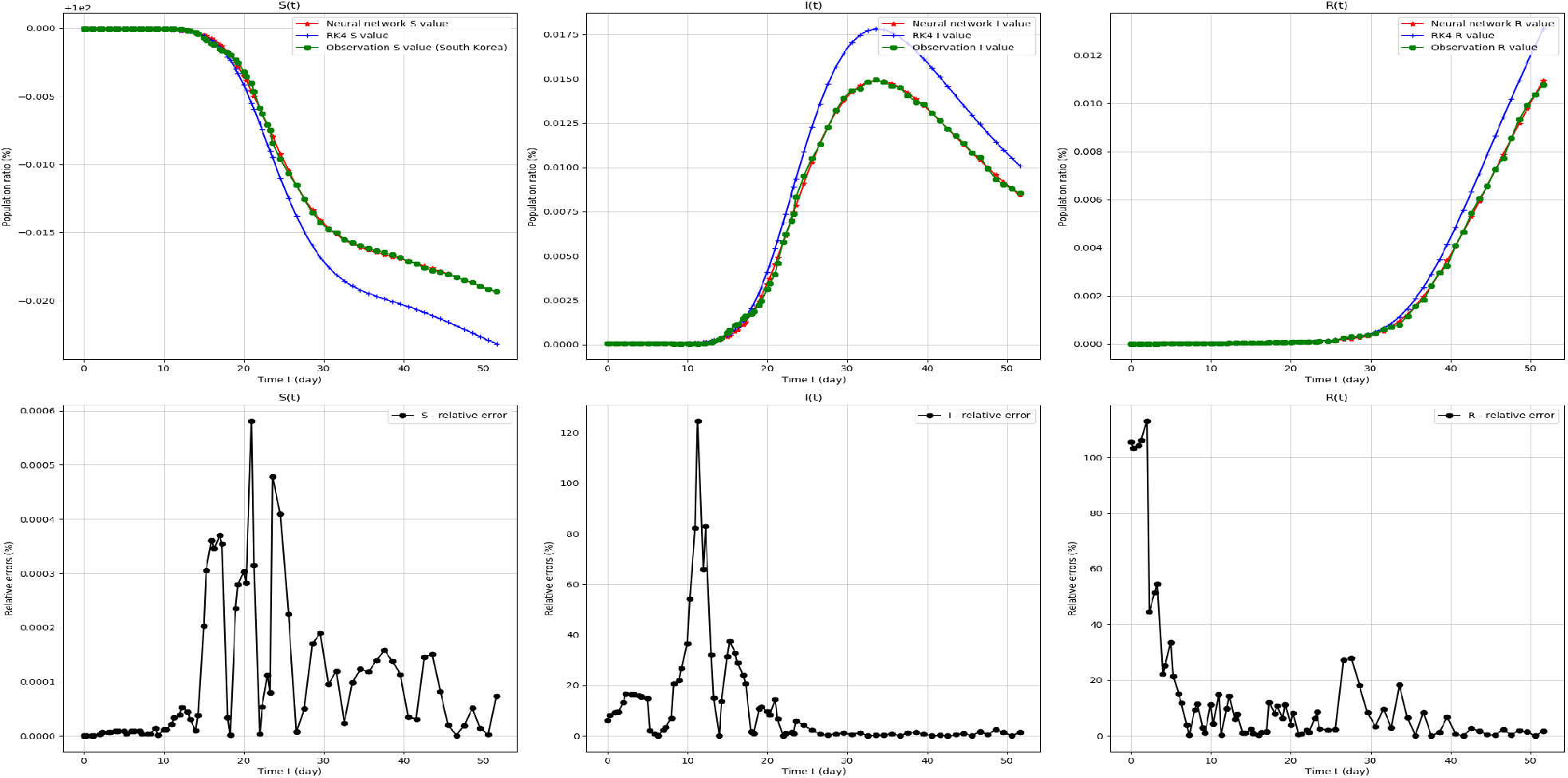
SIR-model target values and relative errors. From 7, February 2020 (*t* = 0) to 30, March 2020 (*t* = 61) in South Korea.

**Figure 8:**
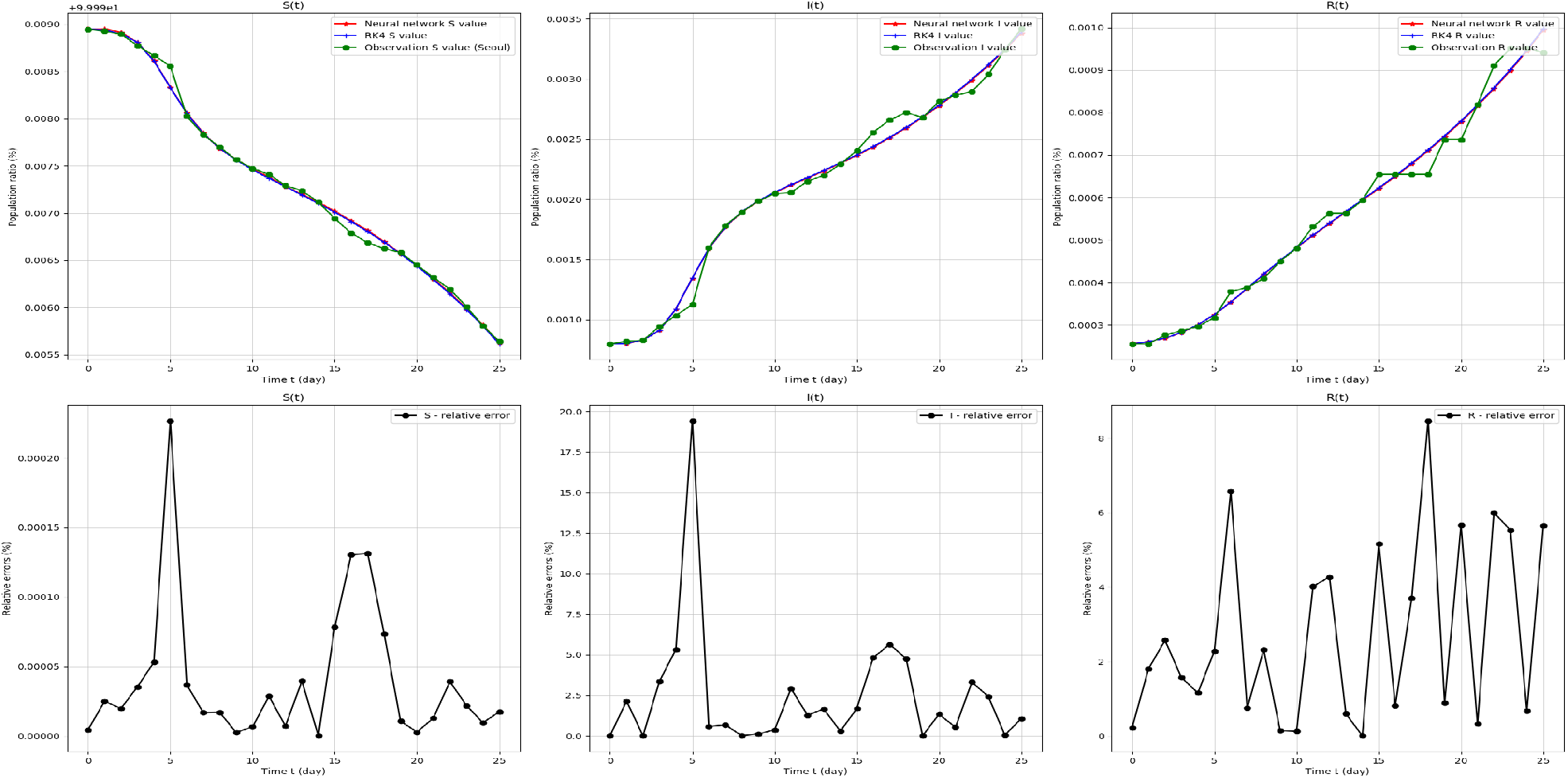
SIR-model target values and relative errors. From 5, March 2020 (*t* = 0) to 30, March 2020 (*t* = 25), in Seoul.

**Figure 9:**
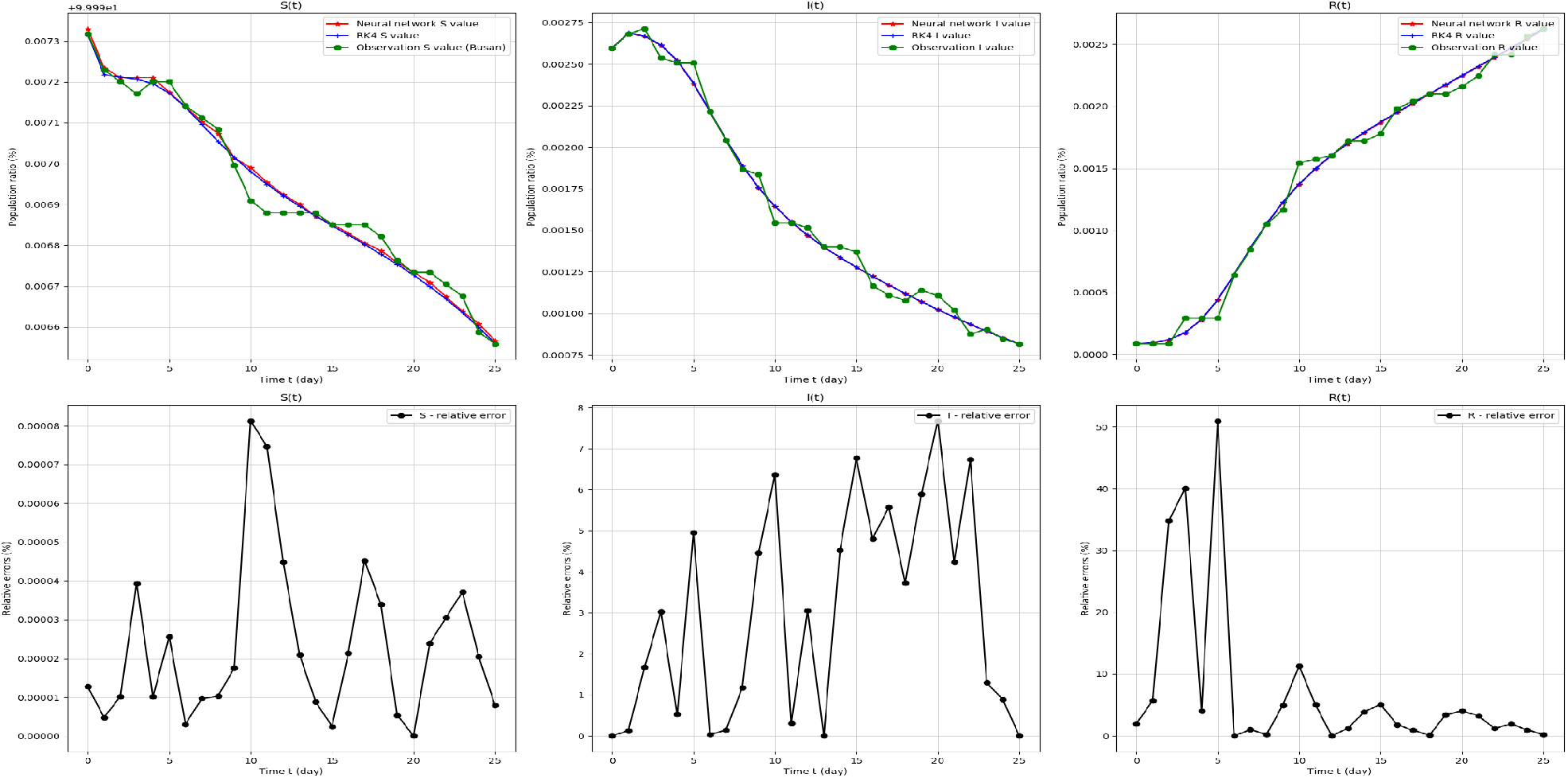
SIR-model target values and relative errors. From 5, March 2020 (*t* = 0) to 30 March 2020 (*t* = 25), in Busan.

**Figure 10:**
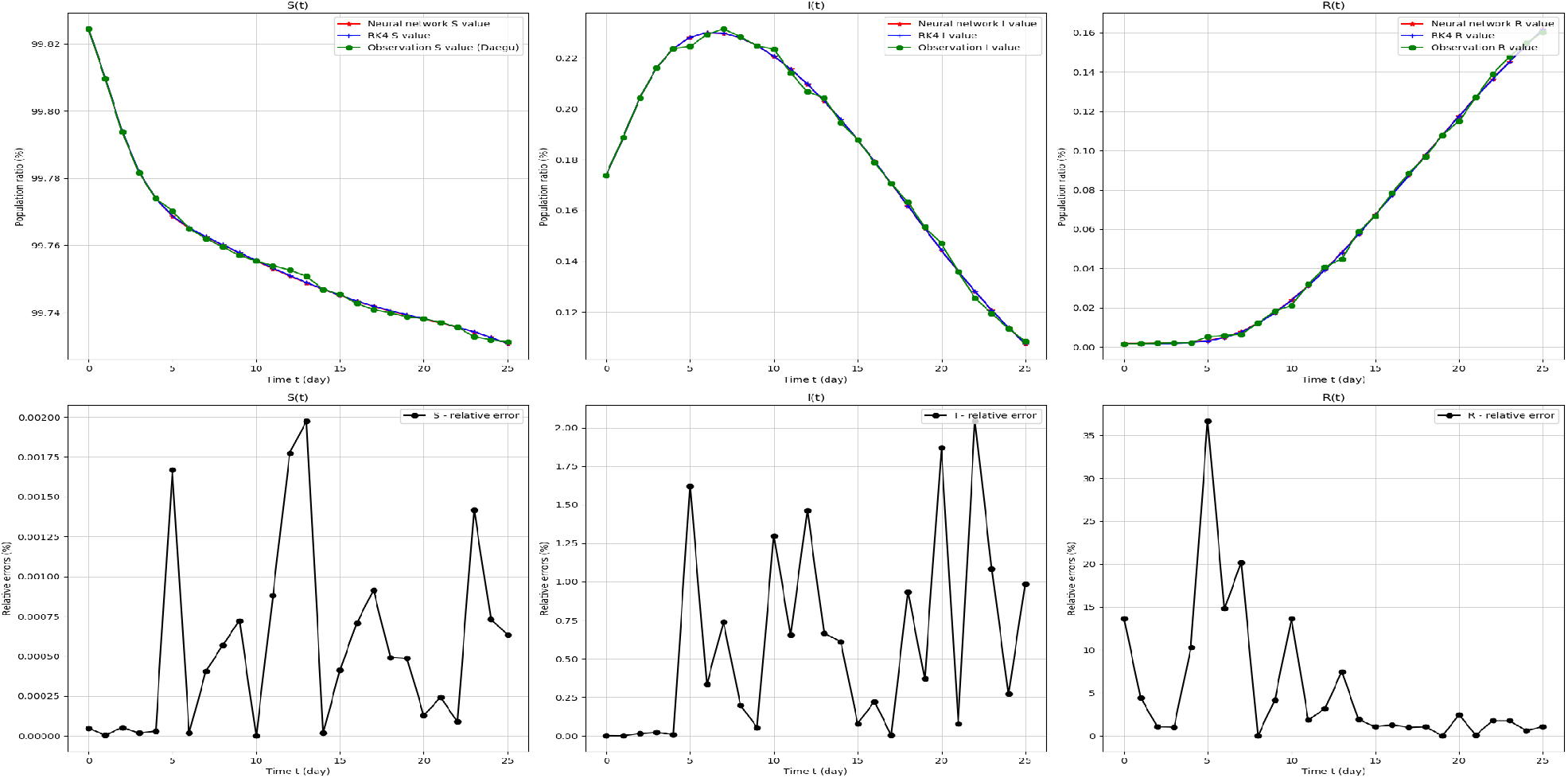
SIR-model target values and relative errors. From 5, March 2020 (*t* = 0) to 30, March 2020 (*t* = 25), in Daegu.

**Figure 11:**
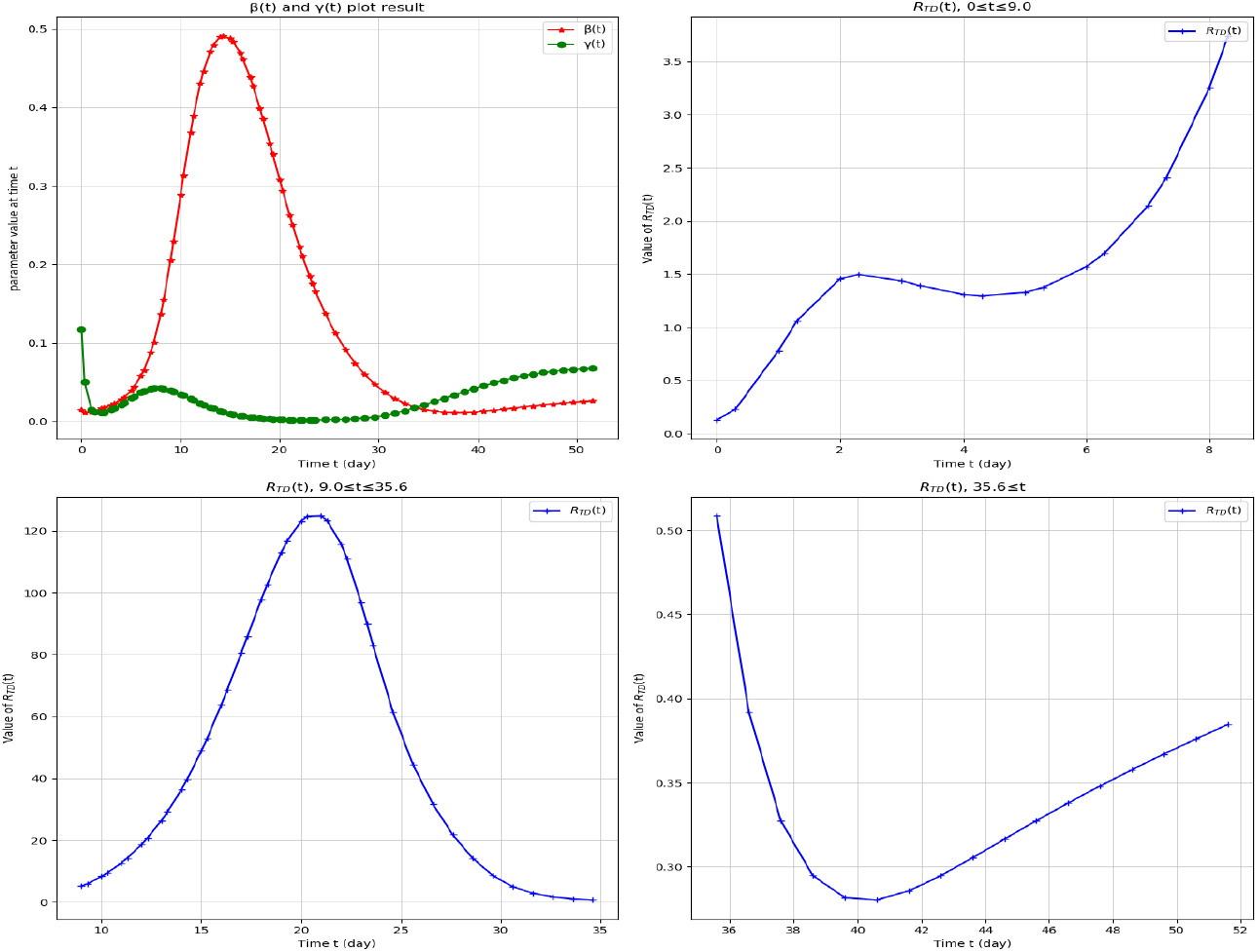
SIR-model Parameter network values and *R*_*T D*_. From 7, February 2020 (*t* = 0) to 30, March 2020 (*t* = 61) in South Korea. We divided the range of *R*_*T D*_ into 3 parts, right top (0 ≤ *t* ≤ 9.0), left bottom (9.0 ≤ *t* ≤ 35.6), and right bottom (35.6 ≤ *t*.

**Figure 12:**
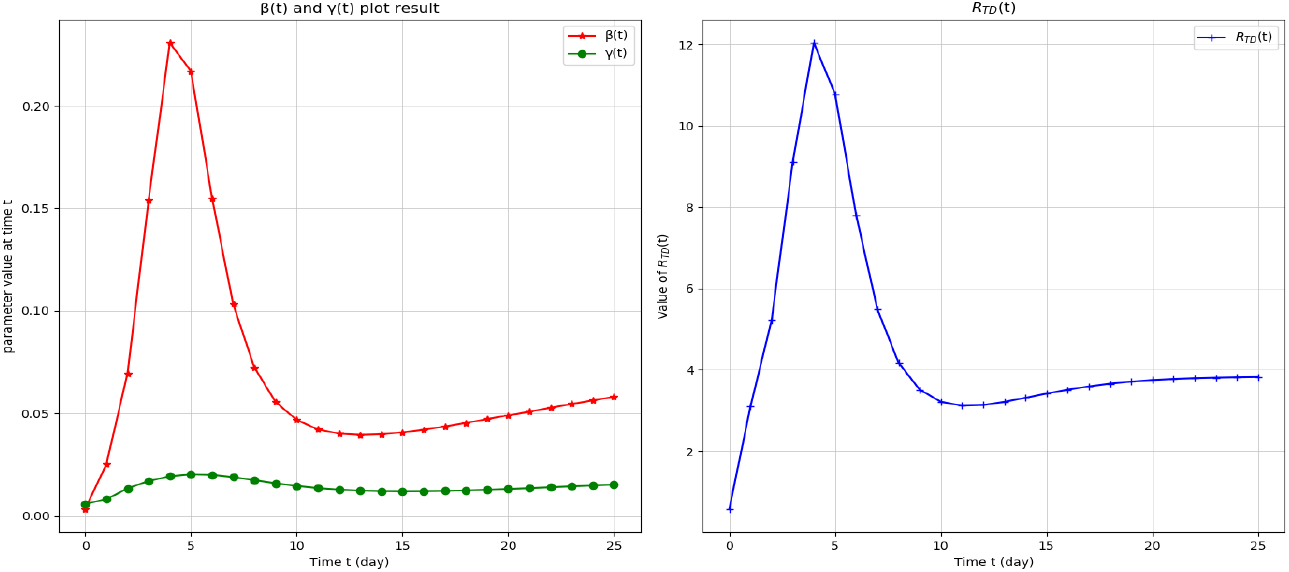
SIR-model Parameter network values and *R*_*T D*_. From 5, March 2020 (*t* = 0) to 30, March 2020 (*t* = 25), in Seoul.

**Figure 13:**
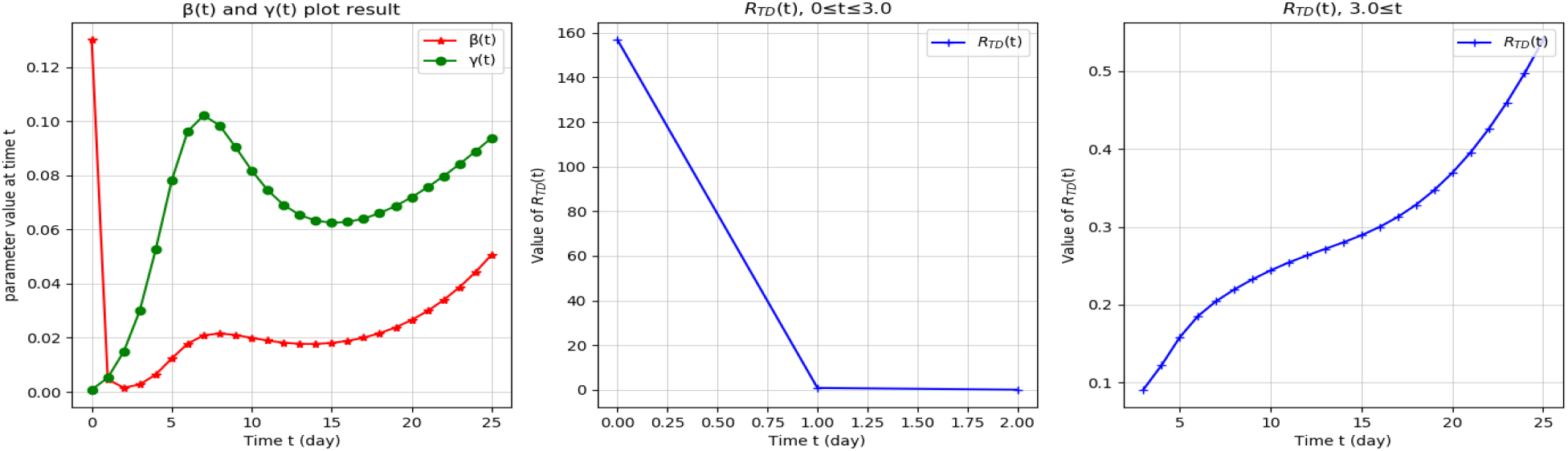
SIR-model Parameter network values and *R*_*T D*_. From 5 March 2020 (*t* = 0) to 30 March 2020 (*t* = 25), in Busan. We divide the range of *R*_*T D*_ into 2 parts: left bottom (0 ≤ *t* ≤ 3.0) and right bottom (3.0 ≤ *t*).

**Figure 14:**
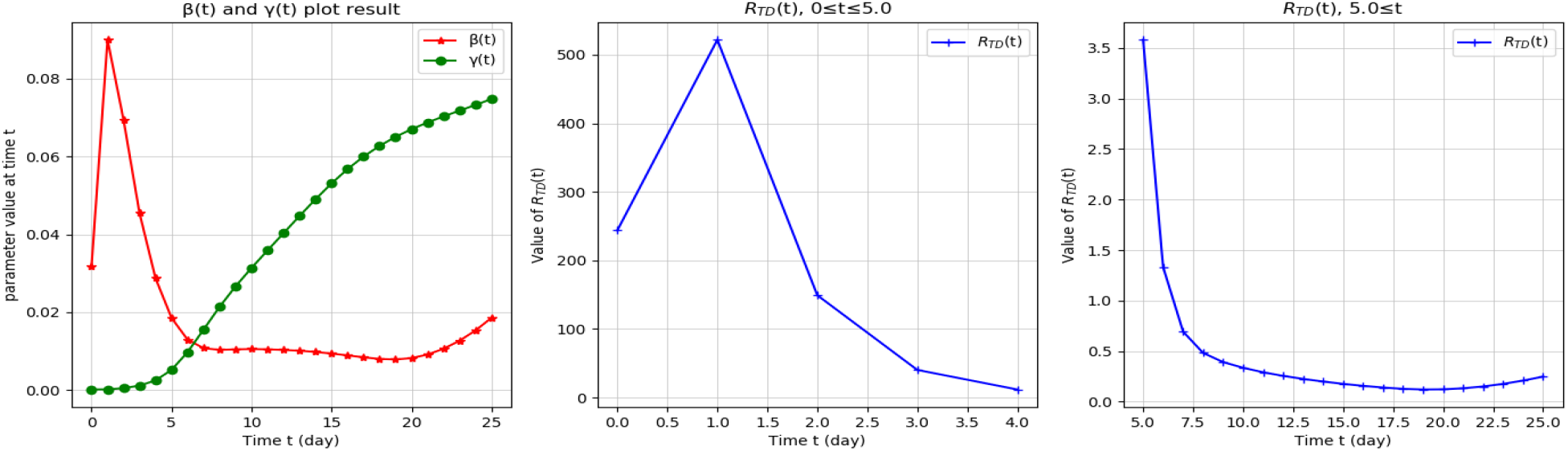
SIR-model Parameter network values and *R*_*T D*_. From 5, March 2020 (*t* = 0) to 30, March 2020 (*t* = 25), in Daegu. We divide the range of *R*_*T D*_ into 2 parts: left bottom (0 ≤ *t* ≤ 5.0) and right bottom (5.0 ≤ *t*).

### 5.1. SIR model results

### 5.2. SIR parameter results

## 6. Discussion and conclusions

Since *R*_*T D*_ is the ratio of *β*(*t*), and *γ*(*t*), *R*_*T D*_ can have an unusually large value when *γ* is small compared to *β*. This situation is observed in the early stage of COVID-19 spread in South Korea (excluding Seoul and Busan,) such as the Shincheonji cult cases. However, following the computation of the basic reproduction number in [15], we obtained the effective reproduction number *R*_*t*_ in the usual range. In the SIR model, we approximated *S ∼* 1, since *S* was large enough compared to *I*. Then, the differential equation 3.5 was approximated by

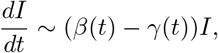

with the solution of this approximated equation being 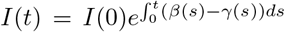, as in the growth model. Therefore, we can consider 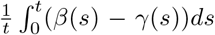 the growth rate, and compute the effective reproduction number by 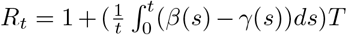 [16, 17], *or* 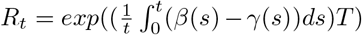 where *T* is the serial interval, as in Figure 15. We used *T* = 4 following [18].

**Figure 15:**
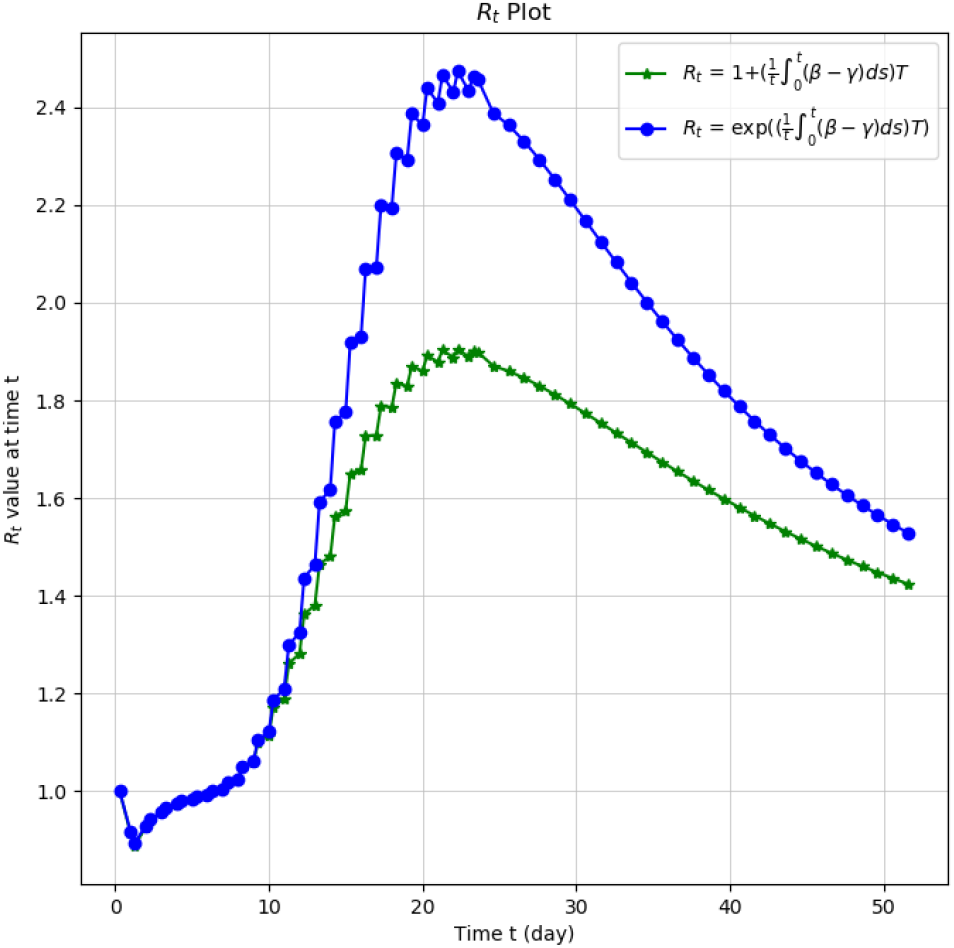
The effective reproduction number.

Using our time-dependent parameters *β*_*net*_, and *γ*_*net*_, we found that traditional numerical algorithms, such as the Runge-Kutta methods, can successfully approximate the SIR model while fitting the COVID-19 data. Moreover, we find that the parameters *β*_*net*_, and *γ*_*net*_ are good enough to fit the model and the data, provided the convergence of the RK4 method to the solution.

At present, 100 years after the Spanish flu, substantial amounts of information are exchanged in real time, with analysis results coming out in real time from all over the world, making the world a gigantic clinical trial. Therefore, it is important for governments to analyze the collected data more rigorously to obtain insights from them, and create sophisticated models that reflect real-time data to guide their policies and curb the COVID-19 spread.

Unlike previous studies, this study was able to model the propagation patterns of COVID-19 more precisely.

## Data Availability

All data is available on this website : http://ncov.mohw.go.kr/

